# Tracing the evolutionary path of the CCR5delta32 deletion via ancient and modern genomes

**DOI:** 10.1101/2023.06.15.23290026

**Authors:** Kirstine Ravn, Leonardo Cobuccio, Rasa Audange Muktupavela, Jonas Meisner, Michael Eriksen Benros, Thorfinn Sand Korneliussen, Martin Sikora, Eske Willerslev, Morten E. Allentoft, Evan K. Irving-Pease, Fernando Racimo, Simon Rasmussen

## Abstract

The chemokine receptor variant CCR5delta32 is linked to HIV-1 infection resistance and other pathological conditions. In European populations, the allele frequency ranges from 10-16%, and its evolution has been extensively debated throughout the years. We provide a detailed perspective of the evolutionary history of the deletion through time and space. We discovered that the CCR5delta32 allele arose on a pre-existing haplotype consisting of 84 variants. Using this information, we developed a haplotype-aware probabilistic model to screen for this deletion across 860 low-coverage ancient genomes and we found evidence that CCR5delta32 arose at least 7,000 years BP, with a likely origin somewhere in the Western Eurasian Steppe region. We further show evidence that the CCR5delta32 haplotype underwent positive selection between 7,000-2,000 BP in Western Eurasia and that the presence of the haplotype in Latin America can be explained by post-Columbian genetic exchanges. Finally, we point to new complex CCR5delta32 genotype-haplotype-phenotype relationships, which demand consideration when targeting the CCR5 receptor for therapeutic strategies.

## Introduction

Humans have been exposed to pathogens over the course of our evolutionary history, and adaptations to them have left numerous signatures in our genomes ^1–3^. In recent years, evidence for selection has been found in genes involved in the development of tolerance against intracellular pathogens and in the inflammatory response against extracellular microbes ^4–6^. These include, for example, the TLR6-TLR1-TLR10 cluster of toll-like receptors, which are crucial components of innate immunity against pathogens, and were likely under positive selection in anatomically modern humans after introgression from archaic hominin groups ^7,8^. More recently, Domínguez-Andrés et al, 2021^9^ showed that alleles associated with cytokine profiles reflecting immune tolerance were under selection during the transition to farming in the Neolithic period, as sedentarism and population density increased, enabling the development of pathogen reservoirs in newly domesticated animals.

Perhaps one of the most intensively debated immune-associated loci previously posited to have been under selection in humans is a 32-bp deletion (CCR5delta32, rs333), which introduces a premature stop codon in the C-C chemokine receptor 5 gene (ENSG00000160791:*CCR5*) ^10–17^. CCR5 is a member of the G-protein-coupled receptor family of proteins and, upon activation, by the CC chemokines CCL3, CCL4, and CCL5, CCR5 and its ligands, plays a critical function in regulating the inflammatory response by facilitating communication between immune cells and the environment ^18–20^. Thus, CCR5 can act as a regulator of the host’s immune response. In 1996, CCR5 was identified as a necessary co-receptor for the macrophage-tropic HIV strains ^21,22^ and it was subsequently reported that CCR5delta32 could provide HIV-1 infection resistance to individuals carrying this allele in homozygous form ^23^. CCR5 is now an important target in preventing and treating HIV infection, using various therapeutic strategies ^24,25^. A female patient with HIV-1 was recently potentially cured for both HIV-1 and acute myeloid leukemia through a CCR5delta32/delta32 haplo-cord transplant ^26^, a method that has successfully cured two similar cases using unrelated donor stem cells with the same genetic modification ^27,28^.

Besides the significant effect on HIV infection, the CCR5delta32 allele has also been associated with other pathological conditions including infection by other viral organisms (including SARS-CoV-2 which causes COVID-19), immune-related diseases, neurological disorders, and various types of cancer ^20,29–41^. Together these studies indicate that the CCR5delta32 allele is pleiotropic and can act as a modulator of a given phenotype expression, with both advantages and disadvantages, depending on the medical context. In this perspective, serious concerns have been raised by the scientific community about possible clinical side effects on “CCR5delta32” CRISPR babies, whose genomes have been edited to confer lifetime HIV immunity ^42–45^.

The evolutionary history of the CCR5delta32 deletion has been widely debated and with conflicting research results ^10,12,13,15–17,46–54^. Today the CCR5delta32 allele frequency (AF) is between 0.10-0.16 in Northern European populations and less than 0.08 in South- and South-East Europe ^12,55^. Outside of Europe the deletion is found only in populations with European ancestry ^56–59^. Past studies have estimated the age of the CCR5delta32 allele with divergent results ranging from ∼700, ∼3,400, and >5,000 years ago ^10–12,15,17^. Positive selection, negative selection, balancing selection, and genetic drift have each been proposed as an explanation for the distribution of present-day gene frequencies ^10,12,13,15–17,46–54,60,61^.

The few studies conducted on the CCR5delta32 deletion in ancient individuals have been constrained by a limited geographic scope and small sample sizes, leading to the possibility of biasing the results by familial relations. So far, the oldest CCRdelta32 alleles have been detected in a 4,900-years-old individual belonging to the Yamnaya culture ^62^ and in several Swedish individuals dating to the Neolithic period (5,250-1,690 BCE) ^48^. However, the latter study raised concerns about allelic dropout during the assaying process, which could lead to genotype misclassification. Two studies conducted on ancient genomes from individuals in central and northern Germany revealed no significant change in the frequency of the CCR5delta32 variant over the past millennium, including during the Black Death pandemic^46,49^. In contrast, a study conducted in Poland reported a nearly doubled frequency of the CCR5delta32 variant from the late medieval period to the present day.^63^

The emergence of paleogenomics has shed light on our understanding of human population history, but evidence from large ancient genomic datasets has been missing in the debate on the CCR5delta32 allele. Due to the degraded nature of ancient DNA, ancient genomic datasets tend to be characterized by short read lengths and post-mortem DNA damage ^64^, impairing the ability to identify indels like the CCR5delta32 deletion. Moreover, mapping efforts in the CCR5delta32 region are particularly challenging because the breakpoint’s flanking regions contain repeated sequences.

In this study, we describe the evolutionary trajectory of the CCR5delta32 deletion as revealed by ancient and present-day genomes. We discovered that the CCR5delta32 allele emerged on a pre-existing haplotype comprising 84 variants, which prompted us to develop a probabilistic model that allows for reliable detection of CCR5delta32 in low-coverage genomes, improving our ability to study the allele’s distribution and impact in space and time.

Applying our model to large ancient genomic datasets (>800 genomes), we find evidence in support of a temporal origin of the CCR5delta32 deletion that is at least 7,000 years BP in age, with a likely spatial origin somewhere in the Western Eurasian Steppe. Furthermore, our findings provide evidence that the CCR5delta32 haplotype underwent positive selection in Eurasia between 7,000-2,000 BP. Analyzing the CCR5delta32 haplotype in individuals from Latin America, we determined that the presence of the allele in this region can be attributed to the Columbian Exchange. This study is the first to provide a comprehensive picture of the CCR5delta32 allele’s evolutionary history across time and space.

## Results

### Identification of three CCR5 haplotypes in Europe

By re-analyzing the European individuals from the 1000 Genomes Project phase 3 (1KGP3) data, we discovered that the CCR5delta32 allele was located on a haplotype with up to 107 variants in high linkage disequilibrium (LD): r^2^ > 0.8 (for details see Table S1). The longest haplotype was identified in the FIN (Finnish in Finland) population, spanning 107 variants including 76 variants with r^2^ = 1. We note that 86 of the 107 variants were also found to be in high LD in the CEU panel (Utah residents with Northern and Western European ancestry), including two variants with r^2^ = 1 (rs113341849 and rs113010081) (Figure 1A). In contrast, in the TSI (Toscani in Italy, r^2^ >0.8, 3 SNPs), IBS (Iberian in Spain, r^2^ >0.8, 3 SNPs), and GBR (British in England and Scotland, r^2^ >0.8, 2 SNPs) panels, we could only identify very few variants in high LD (Table S1). However, these variants were among those with highest LD (r^2^ >0.9) to the deletion in the CEU population. We termed the CEU CCR5delta32 haplotype ‘Haplotype A’ (Figure 1A, Table S1) and identified it in all the 112 CCR5delta32 carriers of the 505 1KGP3 European individuals (AF = 0.111), including three carriers with homologous recombinants of the haplotype (Table S2A).

**Figure 1:**
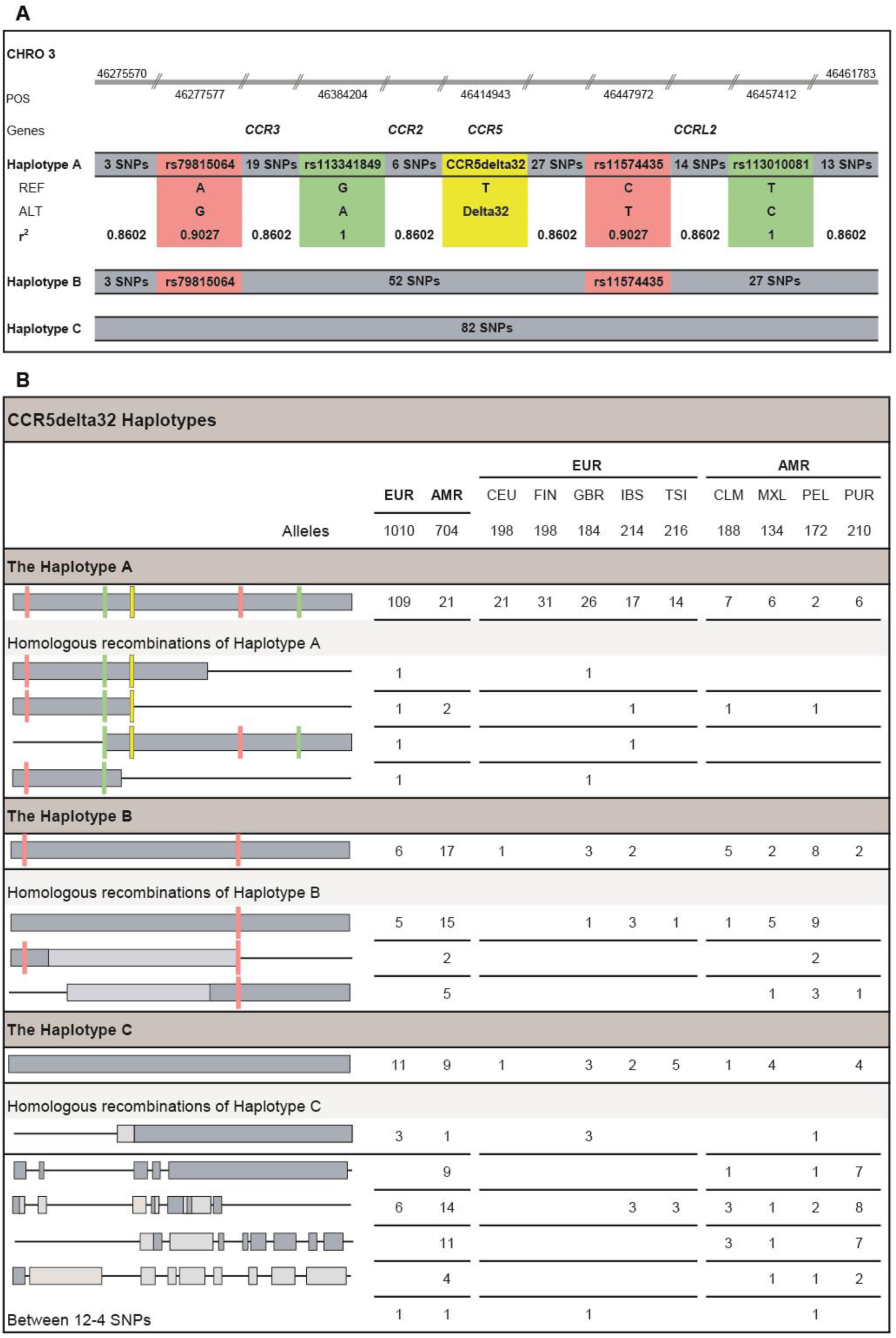
Schematic view of CCR5delta32 and the associated haplotypes: A, B, and C. **A)** Haplotype A consists of CCR5delta32 and 86 tag variants, including two SNPs with an r^2^ value of 1 (rs113341849 and rs113010081, green), two SNPs with an r^2^ value of 0.9027 (rs79815064 and rs1157443, pink), and 82 variants with an r^2^ value of 0.8602 (grey). All r^2^ values are related to the CEU population (**Table S1A**). The haplotype is located on chromosome 3p21.31, spans > 0,19 Mb, and encompasses several genes: *CCR3*, *CCR2*, *CCR5*, and *CCRL2*. Detailed information on the genomic locations of the genes, CCR5delta32, and the 86 tag variants is provided in **Figure S1B**. **B**) Detailed mapping of the Haplotype A, B, and C and their homologous recombinations, among the individuals from 1KGP EUR and AMR populations. The light-gray blocks indicate deviations from different combinations of haplotype blocks. In the Latin American population, a specific homologous recombination of Haplotype A was identified in two individuals from CLM and PUR, which had also been previously detected in an individual from Spain.

Given the strong correlation between the deletion and the variants of Haplotype A in the CEU population, it was surprising that the LD was weaker in Southern and Western Europe (**Figure S1A**). We found that this was caused by another haplotype that included 84 of the 86 tag variants. This haplotype did not include the CCR5delta32 deletion, nor any of the two tag SNPs that were in complete LD (r^2^=1) with the deletion in CEU. We termed this ‘Haplotype B’ (**Figure 1A**) and detected it in 6 of the 505 1KGP3 European individuals (AF = 0.006). Finally, we discovered a third haplotype (‘Haplotype C’) that included 82/84 tag SNPs of Haplotype B in 10 European individuals (AF = 0.009) (**Figure 1A**). Besides these three haplotypes, we identified homologous recombinations and recurrent LD blocks of the two haplotypes without the deletion (Haplotype B and C) (AF = 0.012).

The three haplotypes span >0.18 Mb (chr3:46275570-46461783), including several cytokine receptor genes such as C-C Motif Chemokine Receptor 3, 2, and 5 (*CCR3*, *CCC2*, *CCR5)*, and C-C chemokine receptor-like 2 (*CCRL2)* (**Figure S1B**). To understand the potential functional effect of the variants carried by these haplotypes, we used the Ensembl Variant Effect Predictor (VEP) ^65^, and found that none of the tag SNPs could be annotated with clinical significance, as assigned by ClinVar ^66^. However, from the GWAS catalog ^67^, the tag SNPs with r^2^ > 0.9 have been previously associated with complex traits and diseases, such as Diabetes Mellitus Insulin-Dependent (IDDM), Inflammatory Bowel Disease (IBD), and Alzheimer’s Disease (AD) ^68–70^ (**Table S3A**). Querying the Phenoscanner database ^71,72^ showed that 82 of the 86 tag variants of Haplotype A (including all the tag variants with r^2^ > 0.9) were linked to many of the same phenotypic traits that were already associated with CCR5delta32s’ multiple phenotypes (**Table S3B**). Notably, as the CCR5delta32 deletion is not detectable in traditional SNP-based GWAS analyses, some of these GWAS associations might be caused by the direct linkage to the CCR5delta32 allele.

### Local admixture analysis revealed the European origin of CCR5delta32

We then expanded the analysis to the entire 1KGP3 dataset (2,535 individuals from 26 populations), where we detected 35 individuals having the CCR5delta32 deletion outside of the EUR super-population panel, primarily in populations that have European ancestry (**Table S2A**). In Latin America, we could identify a homologous recombination of Haplotype A in two individuals from CLM (Colombian in Medellin) and PUR (Puerto Rican in Puerto Rico), which we also previously had detected in an individual from Spain (**Figure 1B**). To further investigate local admixture around the CCR5delta32 locus, we applied HaploNet ^73^ to all individuals of the 1KGP3 (**Figure 2A**). Here we found evidence of a European sequence segment in 138 out of 141 individuals who harbored at least one allele of the deletion, while the remaining three individuals from PJL (Punjabi in Lahore, Pakistan) carried insufficient European ancestry proportions for HaploNet to distinguish fine-scale ancestry signals.

**Figure 2:**
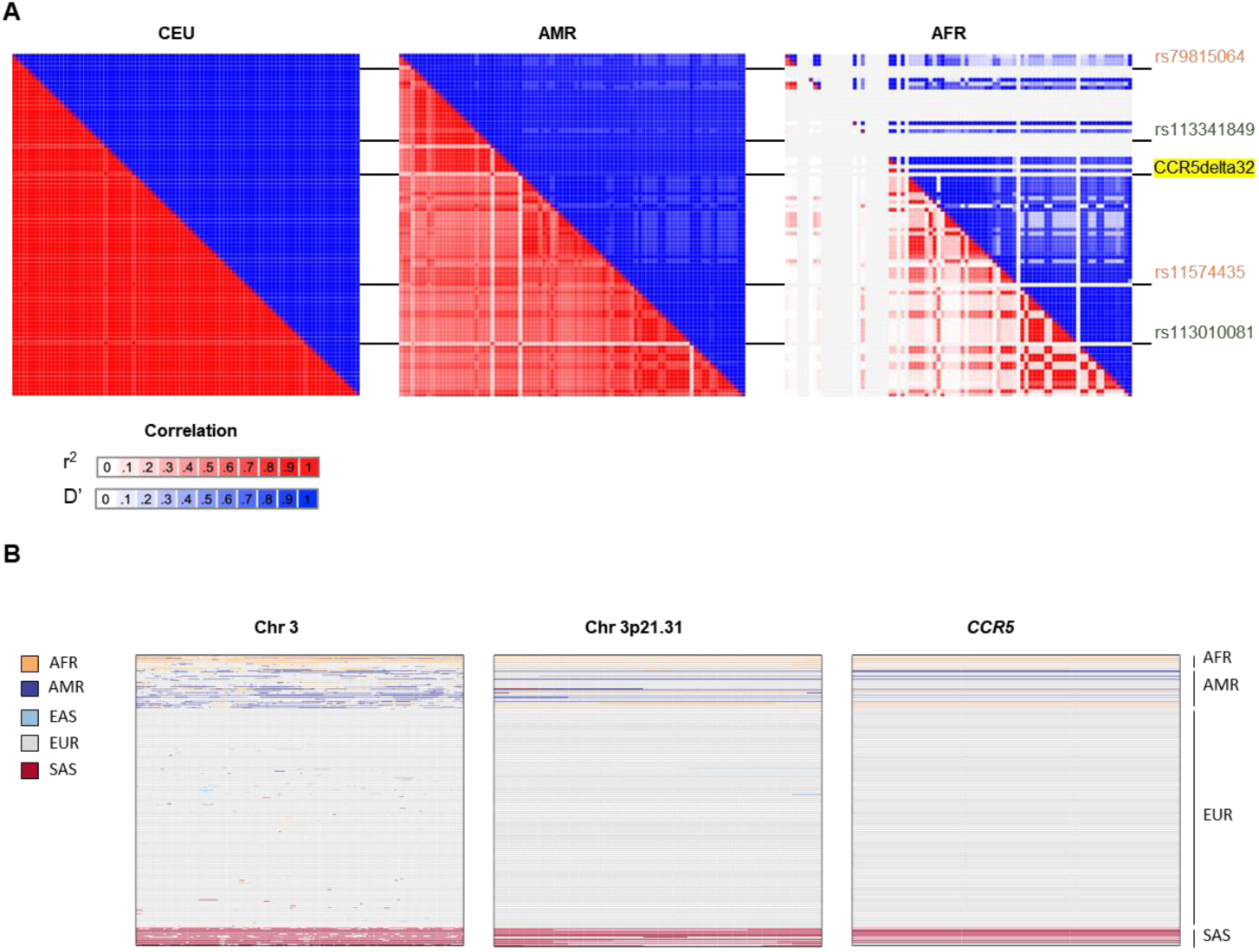
CCR5delta32 locus and Haplotype A patterns of LD in different populations. **A)** Identification of a European CCR5delta32 locus in individuals with the deletion. From the HaploNet analysis, a European sequence segment was identified in 138 out of 141 1KG3 individuals, all genotyped with at least 1 allele of the deletion (**Table S2**). **B)** Heatmap matrices of pairwise LD statistics from Haplotype A in the CEU, AMR, and AFR populations. The strong LD pattern from Haplotype A in the CEU population becomes weaker in Latin America, due to the significantly higher homologous recombination rates we observe from haplotypes B and C in Latin America. These higher recombination rates may be explained by post-Columbian admixture among three groups - African, European, and Native American, as the AFR populations also harbor precursor variations for Haplotype C. The r^2^ values are in shades of red while the D’ values are in shades of blue. Darker values indicate a higher degree of pairwise LD.

Additionally, the complete Haplotype B (AF = 0.024) and shorter homologous recombinants (HR) (AF = 0.031) (**Figure 1B**) were found in significantly higher proportions in Latin Americans than in European populations (chi-square test p-value = 0.002616 and 4.048e-05, respectively). Likewise, we also observed this same pattern for HR Haplotype C (AF=0.057, chi-square test p-value =2.115e-06) (**Figure 1B**). Among the 1,008 individuals with African ancestry originating from the African continent (excluding African Ancestry in SW USA (ASW), and African Caribbean in Barbados (ACB)), we did not detect any of the three haplotypes. We did, however, identify precursor SNPs for Haplotype C (**Figure 2B**). Out of the 82 variants of Haplotype C, 38 had a higher AF in the African population compared to the European population (**Figure S1C**). Therefore, the increased AF of certain haplotype blocks in Latin American populations could be explained by admixture with individuals of African ancestry.

### Haplotype A originated from Haplotype B in Europe

The high frequency of Haplotype A, along with the presence of only four haplotype recombinants (HRs) of Haplotype A in the EUR population (**Figure 1B**), indicates that this haplotype is much younger than Haplotype B and C and or that the CCR5delta32 deletion has been exposed to selection in the EUR population. Hence, based on present-day data alone, this suggests that at some point in the history of present-day Europeans, the two variants rs113341849 and rs113010081 and the CCR5delta32 deletion (all with r^2^ = 1) emerged on Haplotype B, leading to Haplotype A, which is now present at substantial frequencies in present-day Europeans and in certain Latin American individuals, due to post-Columbian admixture (**Figure 2B**).

### A probabilistic framework for calling CCR5delta32 allele in low-coverage aDNA genomes

To trace the evolution of the CCR5delta32 allele through time, we aimed at identifying ancient individuals carrying the deletion. To achieve this, we developed a Haplotype-Aware Probabilistic model for Indels (HAPI), which allowed us to identify the deletion in low-coverage ancient genomes (see Methods). For this, we utilized the information from the four tag SNPs having the highest pairwise LD with the CCR5delta32 allele (r^2^ > 0.90, **Table S1**), as a prior for the presence of the deletion and modeled the information from the reads mapping to the *CCR5* region in the form of a likelihood function. To remove reference bias and improve CCR5delta32 mapping detection, we used both the standard reference sequence and a reference sequence including the deletion, hereinafter referred to as canonical and collapsed references, respectively. We first tested HAPI on 15 genotyped CCR5delta32 genomes from the 1KGP3 and showed that it correctly classified all of them (see Methods). To evaluate HAPI’s performance at lower coverages we created a simulated dataset containing 144 ancient genomes with coverages from 0.3X to 10X (**Figure S2A**). We found that the haplotype-informed prior model performed better compared to a uniform-prior model, with an increase of Matthews Correlation Coefficient (MCC) from 0.79 to 0.97 (**Figure 3A**). Furthermore, we benchmarked the performance of HAPI against the commonly-used GATK HaplotypeCaller ^74^. Here, we found that on the simulated dataset, HAPI could correctly classify 129 genomes out of 144 with an MCC of 0.97, compared to only 79 by the GATK HaplotypeCaller (MCC 0.47), an increase in genomes by 34% (**Figure 3A**). Additionally, it was much more precise on the set that was called by both HAPI and GATK HaplotypeCaller, with MCCs of 0.98 and 0.47, respectively. For the subset of genomes that had coverages between 0.5X and 1X, we found that HAPI could correctly classify 45 of 54 genomes (84%) (**Figure 3B**). For coverage at 0.3X, six of 18 genomes (33%) could be classified by HAPI. Across these very low-coverage genomes (<=1X) HAPI had an MCC of >= 0.84 (**Figure 3B**). For the genomes with coverage lower than 2X, GATK HaplotypeCaller could only correctly classify genomes without CCR5delta32 deletion (**Figure S3A**). The difference between the methods was more pronounced when we stratified by the performance metrics by deletion genotype (RR, RD, DD, i.e. homozygous for the reference, heterozygous, or homozygous for the deletion), where GATK HaplotypeCaller, HAPI with the uniform prior, and HAPI with the informed prior had ROC-AUCs of 0.30, 0.93, and 0.99, respectively (**Figure 3C**). Taken together our model was highly specific for identifying CCR5delta32 allele, even in the heterozygous form and with as little as 0.3X coverage.

**Figure 3:**
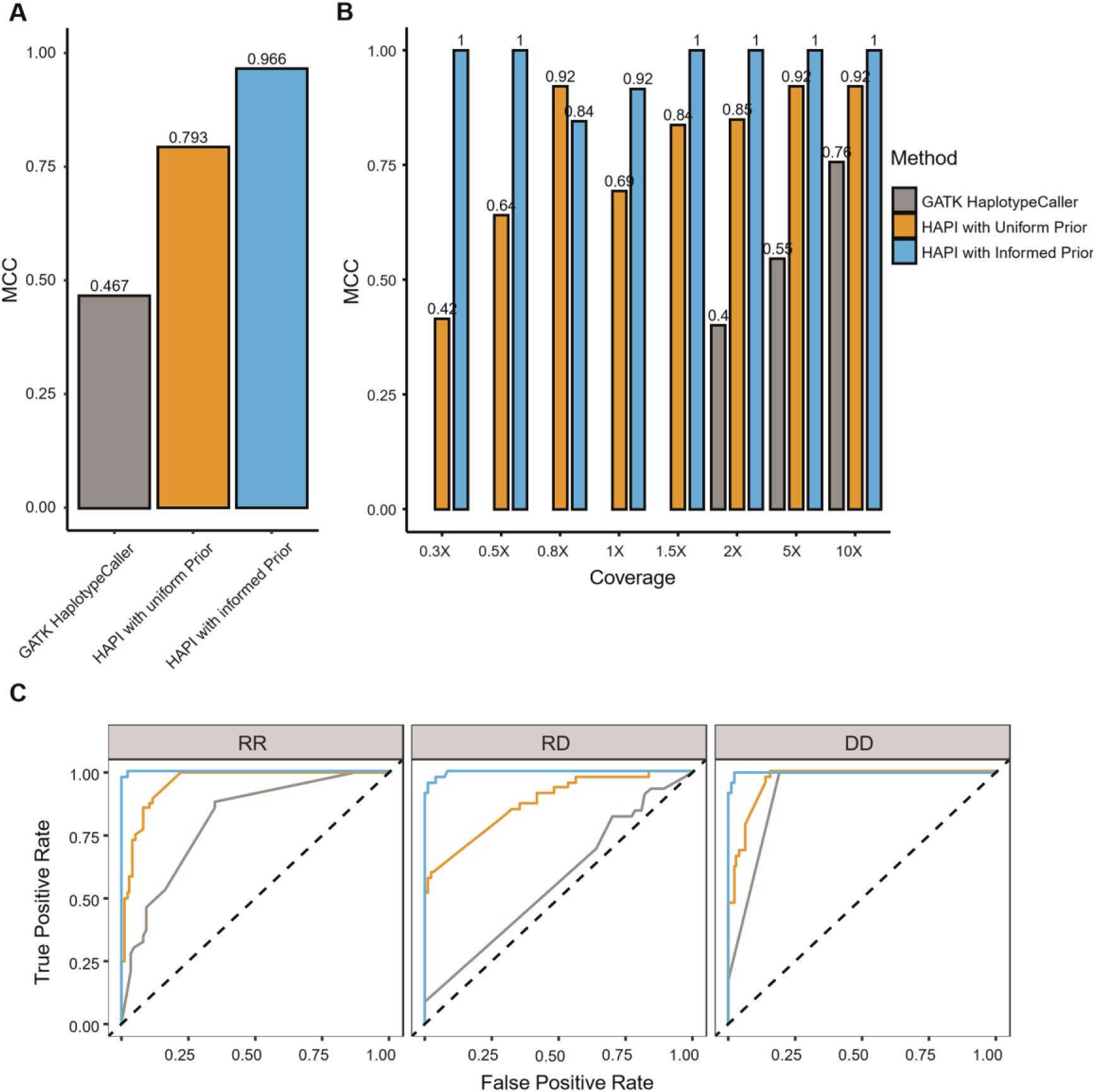
Performances on simulated data. **A)** Comparison of the performance of GATK HaplotypeCaller (grey), HAPI with Uniform Prior (yellow), and HAPI with Informed Prior (blue) on the 144 ancient simulated genomes. Performances are shown as MCC on all the simulated genomes across different coverages. **B)** The same performance comparison but stratified by sequencing coverage. **C)** Performances shown as ROC-AUC on all the simulated genomes stratified by deletion genotype. GATK HaplotypeCaller (grey), HAPI with the Uniform Prior (yellow), and HAPI with the Informed Prior (blue) had ROC-AUCs of 0.30, 0.93, and 0.99, respectively. Further, it is noticeable the precision with which HAPI with the Informed Prior detects the CCR5delta32 allele in heterozygous low-coverage genomes (RD). In these ancient genomes there are often not enough aligned reads to confidently determine the presence of both alleles, which can lead to a biased representation of the genotype in question towards the reference allele. Here HAPI provides a higher degree of reliability and accuracy in genotyping the deletion compared to GATK HaplotypeCaller.

### Applying HAPI to ancient datasets

We then applied HAPI to our extensive ancient DNA dataset, which consisted of 860 genomes^75–77^ from various regions across Eurasia, including a dense sampling collection in northern Europe, specifically in Denmark. The final dataset encompasses consecutive historical eras, ranging from the early Mesolithic and Neolithic periods to the Bronze Age, and extending into the Viking Age. To take into account the complexity of the haplotype and the damaged nature of ancient DNA, we applied two curation steps to the results of the model: a “permissive filter” to reclassify genomes that had artifacts typical of ancient DNA damage, and a “strict filter” to reclassify genomes which were likely harboring the Haplotype B (see Methods). Across the ancient DNA dataset, we found that 418 genomes had at least one read mapping to the *CCR5* region from either the canonical or collapsed reference and having at least 6 bases overlapping the CCR5delta32 breakpoint. Using this approach, we identified the CCR5delta32 allele in 43 and 38 individuals using the permissive and strict filters, respectively (**Figure 4**, **Table S4**). From the Allentoft et al. (2022)^77^ dataset, spanning the Mesolithic and Neolithic, four individuals were identified with the deletion using the strict filter and four individuals were classified with Haplotype B across the different output schemes from HAPI (**Table S4**). Only 31 out of 101 genomes from the Bronze Age data^76^ met the criteria for the analysis by HAPI and, although the sample pool was small, we detected one sample with the CCR5delta32 deletion using the strict filter, and one sample carrying Haplotype B (**Table S4**). From the Viking dataset ^78^, 252 of 442 genomes passed the HAPI inclusion criteria (**Table S4**). From these, 33 genomes were detected to have the CCR5delta32 deletion with the strict filter (Haplotype A, AF = 0.065) and two genomes were identified as having Haplotype B (AF = 0.003). Furthermore, we observed 21 genomes having portions of Haplotype C (>20 proxy SNPs). Detailed view of the location of the ancient DNA genomes is provided in **Figure 4** and **Table S4A-C**.

**Figure 4:**
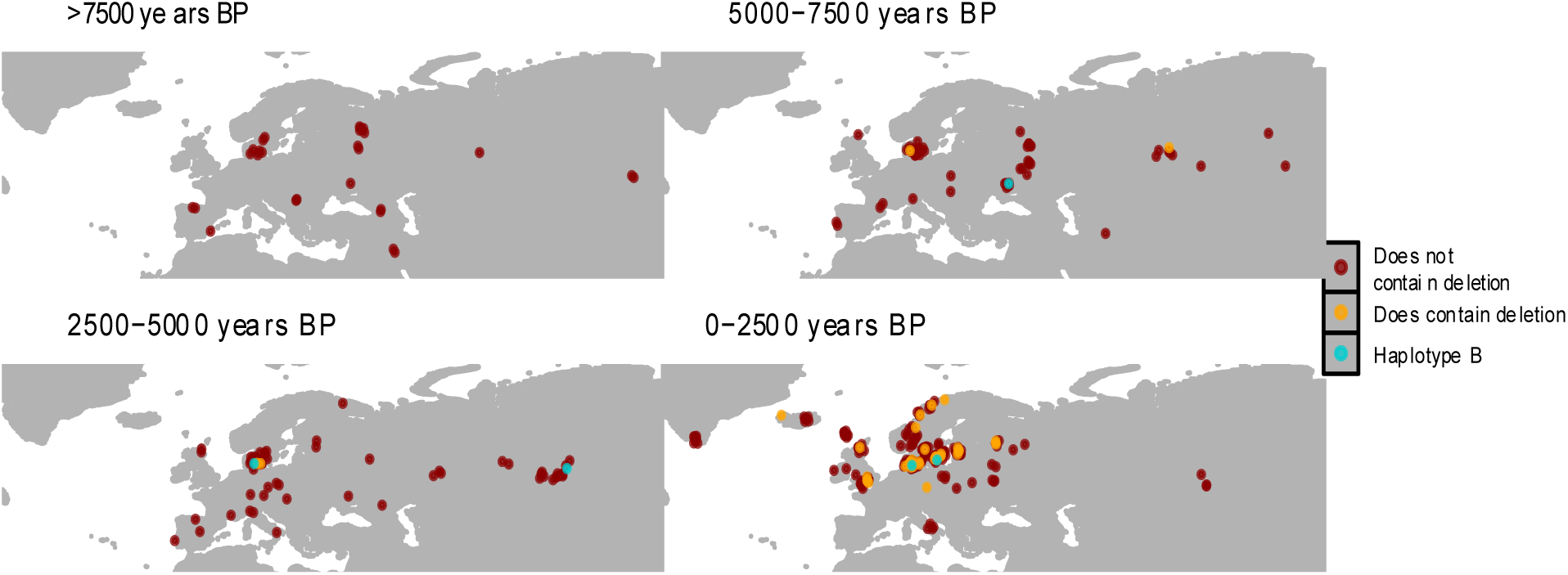
Geographical locations of the ancient genomes genotyped for the CCR5delta32. Map of distribution of ancient genomes genotyped with the permissive filter, faceted by four time periods and colored based on the presence (yellow) or absence (red) of the CCR5delta32. Blue dots correspond to genomes that are classified as having the deletion in the permissive filter genotype call set, but not having the deletion according to the strict filter (The affected genomes are NEO300, NEO590, RISE509, VK316, VK342, see **Table S4**).

### Haplotype A and B were present in Denmark more than 6,000 years ago

In our Mesolithic and Neolithic dataset^77^ we had an extensive collection of ancient genomes from Danish individuals, totaling 100 genomes. We found evidence that the CCR5delta32 allele (Haplotype A) was present in Denmark over 6,000 years ago (NEO855: 6299 cal. BP), as well as in an individual carrying the Haplotype B (NEO683: 7521 cal. BP) (**Figure S4A)**. This places the CCR5delta32 allele in the Danish Ertebølle culture, a hunter-gatherer and fisher, pottery-making culture, dating to the end of the Mesolithic period ^79^. Both genomes were identified to be of the Western Hunter-gatherers ancestry group (HG_EuropeW), which were the predominant ancestry group in Denmark at the time ^77^. In contrast, the Danish individuals detected with the Haplotypes A and B in the later Neolithic and early Bronze Age harbored Steppe-related ancestry (EUR_BA). During the transition from hunter-gatherer to Neolithic and Bronze Age periods, Denmark’s population genomic landscape underwent significant changes^77^. These changes involved the replacement of hunter-gatherer populations and the introduction of Steppe-related ancestry during the late Neolithic and Bronze Age periods.

### Haplotypes A, B, and C were present in Mesolithic and Neolithic Periods across Eurasia

Outside of Denmark, we detected the CCR5delta32 allele in Russia (NEO309: 5824 cal. BP) as well as Haplotype B in Ukraine (NEO300: 6678 cal. BP), Sweden (NEO27: 9693) and Portugal (NEO631: 7135 cal. BP). Interestingly, we detected nine genomes having between 19 and 69 variants from the Haplotype C in Russia (Table S4 B and C), with the oldest sample dating 10,853 cal. BP (NEO202: 69/82 variants). Further, a sample (NEO646) from northwest of Spain dated 8,274 cal. BP was also detected with 35 variants from Haplotype C. Together these results show that, although there was a deep genetic divide between the western and the eastern Eurasia populations ^77^, both groups carried fragments of the three haplotypes (**Figure S4**, Data provided in **Table S4**).

### Evidence for ancient selection operating on the deletion

Based on these results, we aimed to model the spatiotemporal frequency dynamics of the CCR5delta32 allele across West Eurasia, to reconstruct the evolutionary history of this allele and investigate the evidence in favor of positive selection at the locus. We used a modified version of CLUES^80,81^ to infer allele frequency trajectories over time using ancient genomes. In addition to our different genotype call set (strict filter and permissive filter), we evaluated the trajectories if conditioned on allele frequencies observed in present day European populations (**Figure 5**, **Figure S5**). Across these analyses, we observed a rapid rise in the CCR5delta32 frequency until 2,000 years BP, followed by a stabilization of the frequency until the present. When using the strict filter call set and modern ascertainment (**Figure 5B, Figure S5**), however, we observed a very recent uptick in frequency. This was likely an artifact of under-calling of the ancient genomes under this filtering scheme, causing the model to reach present-day frequencies very quickly. We found significant evidence for positive selection acting on the CCR5delta32 allele in the ancient past using both strict and permissive filters against a neutral model (with p-values of 2.27e-3 and 9.34e-3, respectively). In addition, when conditioning on present-day frequency, we obtained even greater significance levels in favor of selection (with P values of 2.69e-8 and 3.41e-5 for strict and permissive filters, respectively. We estimated that a large selection coefficient (s > 0.01) better explained the initial allele frequency rise. When using the strict filter calls, the coefficient was predicted to be smaller (s = 0.0198 with conditioning, s = 0.0152 without conditioning) than when using the “permissive filter” deletion calls (s = 0.032 with conditioning, s = 0.0208 without conditioning). The best posterior estimates for the age of the CCR5delta32 deletion from the CLUES analysis were 9,128 and 7,714 years BP. **Table S5** provides a detailed view of the results.

**Figure 5:**
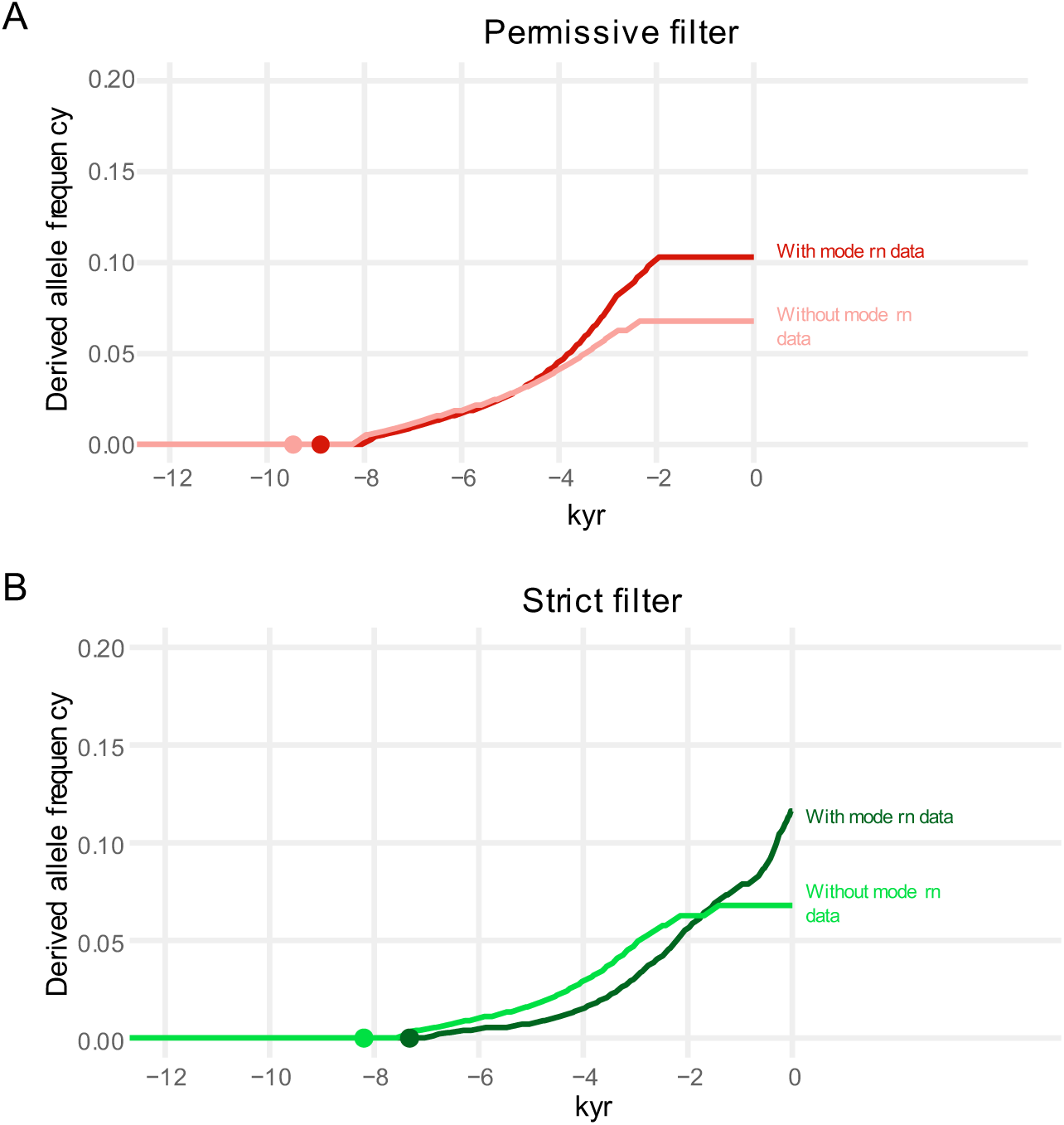
CCR5delta32 allele frequency trajectory. Maximum likelihood trajectory of the CCR5delta32 estimated using CLUES. **A)** Results obtained using permissive filter. **B)** Results obtained using the strict filter. The dots in each figure represent the age estimate of the variant either with or without conditioning on modern ascertainment.

### Spatiotemporal allele frequency dynamics

To investigate the spread of the allele, we fitted a two-dimensional diffusion-advection method that integrates present-day and ancient human genomes to infer allele frequency dynamics across space and time ^82^. The method infers parameters associated with how fast the allele spreads across the landscape and how fast it increases in frequency locally due to positive selection. It also estimates the likely geographic origin of the allele, given the data. Because the CLUES analysis indicated that the allele frequency dynamics changed before and after 2,000 years BP, we partitioned our spatial inference framework into these two periods, allowing the method to find two separate selection coefficients and diffusion parameters for each period (**Figure 6 and Table S6**). We inferred the allele origin to be in the Western Eurasian Steppe region using both filtering schemes, with the strict filter placing the allele more Eastwards compared to the permissive filter. This was followed by a rapid longitudinal expansion in the earlier time period. Regardless of the call set, the selection coefficient was inferred to be higher in the time period before 2,000 years BP (**Figure 6B**), consistent with the CLUES analysis (**Table S5**), suggesting that selection likely operated early in the history of the allele (i.e., during the late Neolithic and Bronze Age). The selection coefficient estimate was higher using the “permissive filter” than when we used the strict filter, likely due to the younger allele age estimate (**Table S6**).

**Figure 6:**
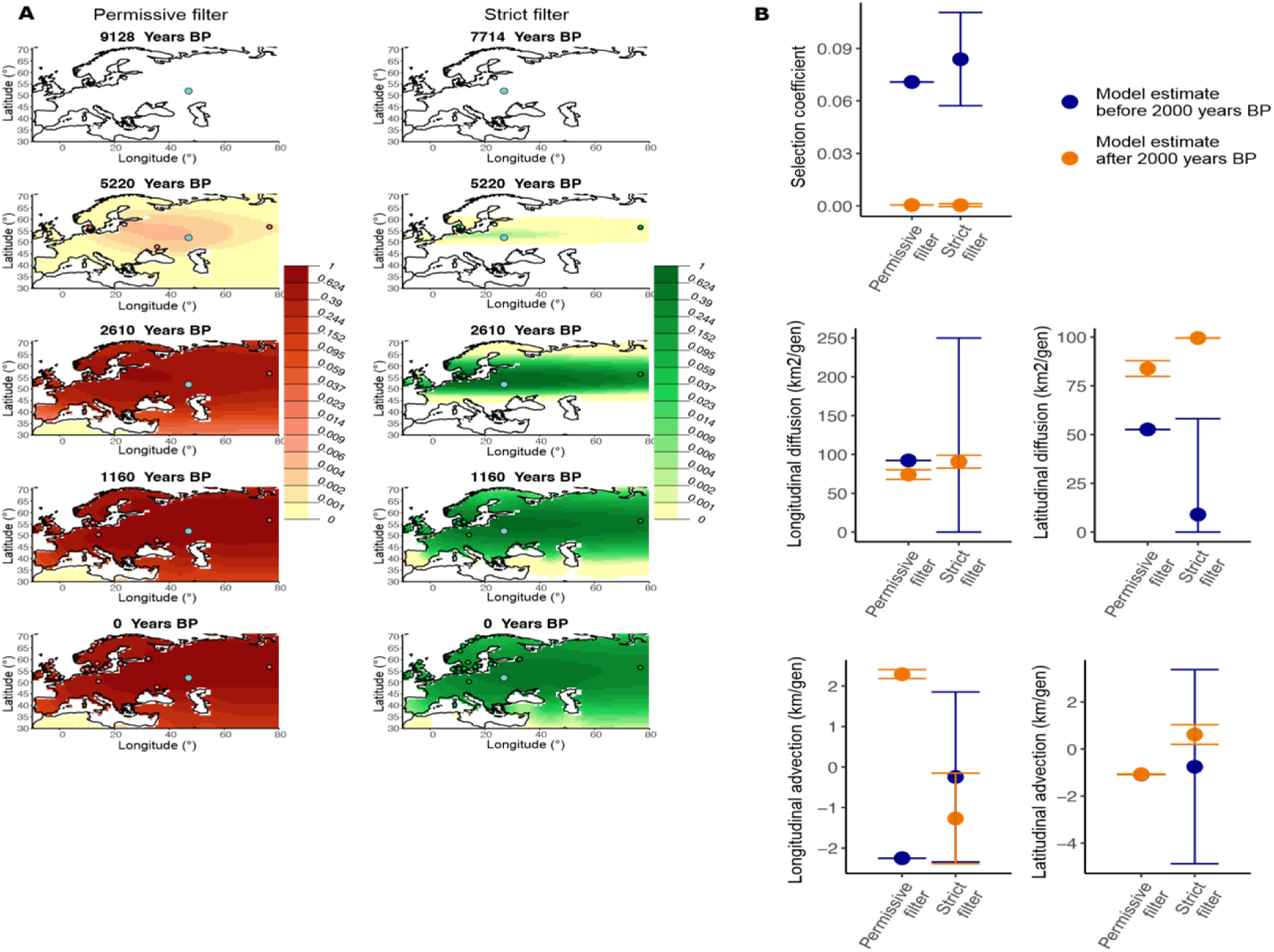
CCR5delta32 allele frequency dynamics across West Eurasia. **A)** Spatial allele frequency dynamics inferred by the diffusion-advection method. Left - permissive filter, right - strict filter. The green and red dots are genomes containing the deletion that are at least as old as the year indicated in each corresponding time slice. The light blue dot corresponds to the inferred origin of the allele. **B)** Parameter estimates from the spatiotemporal diffusion analysis used to generate allele frequency dynamic maps along with 95% confidence intervals. Results are shown for permissive and strict filter genotype call sets for time periods before and after 2000 years before present. The selection coefficient estimates indicate that selection likely operated early in the history of the allele, during the late Neolithic and Bronze Age.

## Discussion

This study provides fundamental new insights into the evolutionary history of the CCR5delta32 allele. Our discovery and mapping of the Haplotypes A, B, and C in present-day genomes led us to develop a probabilistic model, HAPI, to investigate the CCR5delta32 allele in ancient genomes. The model allowed us to reliably detect CCR5delta32 allele in genomes with as little as 0.3X coverage. Based on this, we date the deletion to be at least 7,000 years BP in age, possibly arising among peoples occupying the Western Eurasian steppe region in the Neolithic. We also show that the CCR5delta32 allele was exposed to positive selection during the late Neolithic and Bronze Age, followed by stability in the AF until the present day.

Applying the knowledge of Haplotype A, combined with the evidence from HaploNet, we can now confirm earlier studies’ presumption of a European origin of the CCR5delta32 allele^12,55^. The Columbian Exchange, which was considered to have facilitated genetic admixture among three groups – African, European, and Native American ^83,84^, can account for the significantly higher recombination rates we observe from Haplotype B and C in Latin America compared to European populations, along with the higher AF from some of the variants including in haplotype C (**Figure 2B, Figure S1C**). Thus, we can propose to include the CCR5delta32 allele, together with the variants rs113341849 and rs113010081, as European ancestry-informative markers. Furthermore, the CCR5delta32 genotype can be reliably imputed from SNP arrays using the two r^2^=1 tag SNPs (rs113341849 and rs113010081), as they are located on each side of the CCR5delta32 allele and therefore will encounter most possible recombinations of the Haplotype A.

Previous studies investigating the evolutionary history of the CCR5delta32 allele have been either restricted to contemporary individuals ^12^ or used very few ancient genomes from limited geographic areas ^46,49,63^. Here we present results obtained using a large comprehensive set of ancient genomes (>800 genomes) combined with modern genomes. The CLUES analysis revealed that the allele rose quickly in frequency in the period before 2,000 years BP, followed by a period of AF stabilization, over which the allele remained at around 10% frequency from 2,000 BP onwards. This agreed with findings from Bouwman et al. 2017^46^ and Hummel et al. 2005^49^, which posited a period of recent allele stability over the past millennium in Central and North Germany. Based on our data, the allele had an origin in the Western steppe and a fast rapid diffusion eastwards and westwards early in its history, partly coinciding with the eastward movements from the Steppe during the Bronze Age^76,85^. We note, though, that the origin of the allele inferred by the model is highly dependent on the first instances of the allele in the data, and thus is highly dependent on the mode of deletion calling. Under the curated calling schemes, the lower inferred counts of the allele during the Mesolithic and Neolithic lead the model to estimate a fast longitudinal diffusion, as the most likely allele frequency surface rapidly shifts from complete absence to widespread presence of the allele in distant regions across the continent. The rapid longitudinal spread of the allele is consistent with previous evidence for long-distance dispersal of the allele ^12^ though our ancient data suggests this dispersal occurred earlier than previously thought. Our estimated age of the allele is consistent with a more ancient origin as postulated in Sabeti et al 2005^15^, rather than a recent origin as suggested in other studies ^11,17^. All age estimates we obtained were older than 7,000 years BP (posterior estimates 9,128 and 7,714 years BP).

We found significant evidence of positive selection acting on the CCR5delta32 allele in the ancient past, when fitting the data to the CLUES model. When we conditioned the CLUES trajectories on reaching the frequencies observed in present-day data, they result in stronger rises in frequency compared to using ancient data alone, which in turn results in more significant p-values in the rejection of neutrality. This likely indicates an undercounting of the allele in the more ancient time periods. Regardless of the calling scheme, we found significantly large selection coefficients when deploying the spatiotemporal spread model, particularly in the early time period, but no evidence for selection after 2,000 years BP. Of note, however, is that the spatiotemporal model is deterministic, and thus necessarily underestimates the amount of allele frequency stochasticity that occurs during the period under study, so the selection coefficient inferred under this model may be an overestimate. Very recently, Le and colleagues, 2022^61^, found no evidence for the selection of the CCR5delta32 allele during ancient times. However, that result was obtained using a CCR5delta32 proxy SNP, rs73833033, that we found to have r^2^ < 0.8 and therefore the proxy SNP was not included in Haplotype A. Their analysis therefore did not adequately count CCR5delta32 alleles.

The notable increase in CCR5delta32 allele frequency prior to the Iron Age implies that the high frequencies of this allele in modern-day Europe cannot be caused by Medieval Plague as hypothesized previously^46,49^. Instead, the selection signature may have resulted from pressures exerted by previous outbreaks or other pathogens that existed in the past ^14,16,86^. The observed spread was also not consistent with the Viking-spread hypothesis ^50^. Likewise, our age estimation of the CCR5delta32 allele does not support this hypothesis. Instead, the rapid longitudinal spread that we infer (approx. 60-100 km^2 per generation, **Table S6**) and the rapid rise in frequency observed during the Bronze Age suggests a possible spread associated with the Late Neolithic and Early Bronze Age expansion of steppe-related ancestry into Europe ^76,85^.

Today, immunological genetic signatures by selection and/or adaptation through admixture can be observed in the human genome ^2,3^. Our data shows that the CCR5delta32 (Haplotype A), could very well be among these genetic signatures. We cannot point out a direct cause for the increase of CCR5delta32’s allele fre uency during the Neolithic and early Bronze Age, but it is clear that Haplotype B did not undergo the same evolutionary trajectory. The key to understanding the driving forces for the CCR5delta32 deletion is challenged by immune system redundancy and the immune gene pleiotropy ^87,88^. A hypothesis could be that the CCR5delta32 allele with the 86 tag variants is associated with cytokine/chemokine profiles reflecting immune tolerance, which has been shown to be under selection during the Neolithic age ^9^.

Finally, the fact that individuals bearing the CCR5delta32 allele also harbor a defined haplotype widens the complexity of the deletion effects. The CCR5delta32 deletion has been studied extensively for more than two decades, especially due to its strong link to HIV-1 infection resistance and thereby the potential to target CCR5 for HIV treatment and for HIV pre/post-exposure prophylaxis medicine ^25,27,89,90^. These therapeutic approaches include gene-editing techniques like CRISPR, CCR5 blockade using antibodies or antagonists, or combinations of both ^25,91^. Further, the CCR5delta32 allele can be viewed as a pleiotropic variant, due to its influence on multiple phenotypic traits, e.g. autoimmune and inflammatory diseases, cardiovascular diseases, neurodegenerative disease, and cancer ^20,29,32,44,92^. It is possible that some of the CCR5delta32 tag SNPs contribute to the pleiotropic nature of CCR5delta32, although *in silico* analysis shows no direct clinical significance. More precisely, the gene expression of cytokine receptors (*CCR3*, *CCR2*) and *CCRL2* might be affected by one or more of the tag SNPs, leading to modulation of chemokine-chemokine receptor signal transduction ^19,92,93^. This calls for further studies to elucidate these possible direct or indirect effects. Thus, the tag SNPs should be considered when analyzing causes of the CCR5delta32 pleiotropic effects and when developing therapeutic approaches, based on mimicking the naturally occurring CCR5delta32 genotype-phenotype correlations. Therefore, our results point in a direction of new complex CCR5delta32 genotype-haplotype-phenotype relationships, which demand consideration when targeting the CCR5 receptor for therapeutic strategy.

### Limitations of the study

We have striven to evaluate our results in light of the challenges encompassed by ancient datasets, such as DNA damage sequencing profiles, low-coverage genomes, and patchy spatiotemporal sampling. We developed and applied the HAPI model with awareness of the three different CCR5 haplotypes. By applying different filter schemes to the CCR5delta32 classification of the ancient genomes, we were able to inspect the impact on the ancient sample sizes. Despite these considerations, our results need to be verified through further genome sampling, especially from the European Neolithic period.

## Supporting information

SI_CCR5delta32_Tables.xlsx

## Data Availability

All datasets used in our study are available online, except for the Meso/Neo dataset (/doi.org/10.1101/2022.05.04.490594), which is soon to be published on The European Nucleotide Archive (ENA).

https://www.ebi.ac.uk/ena/browser/view/PRJEB9021

https://www.ebi.ac.uk/ena/browser/view/PRJEB37976

http://www.1000genomes.org/

https://doi.org/10.1101/2022.05.04.490594

## Acknowledgments

K.R., L.C. and S.R. were supported by the Novo Nordisk Foundation (grant NNF14CC0001 and NNF21SA0072102). E.K.I.P was supported by the Lundbeck Foundation (grant R302-2018-2155) and the Novo Nordisk Foundation (grant NNF18SA0035006). E.K.I.P. and R.A.M. were additionally supported by a Villum Young Investigator grant given to F.R (project no. 00025300).

## Author contributions

Conceptualization: K.R., L.C., M.E.A., E.W., F.R., and S.R.; Methodology: K.R., L.C., and S.R.; Data curation: M.S., M.E.A., and E.W.; Investigation: K.R., L.C., R.A.M., J.M., and E.K.I-P.; Software: K.R., L.C., R.A.M., J.M.,T.S.K., and E.K.I-P.; Formal Analysis: M.S.; Writing – original draft: K.R., L.C., R.A.M., F.R., and S.R.; Writing – review and editing: all authors.; Resources: E.W. and M.E.A.; Supervision: F.R. and S.R.

## Supplemental Files

**Supplementary Figures 1-6**

**Figure S1:** Details information of Haplotype A: LD statistics, genomic location and the AF of the proxy variants.

**Figure S2:** Workflow of the data analysis on the simulated ancient samples and details of the overlapping lengths.

**Figure S3:** Assessing GATK HaplotypeCaller, Mismatch Rates, and HAPI performance at different overlapping lengths.

**Figure S4:** Ancient and present sample distribution. **Figure S5:** Allele frequency trajectory inferred by CLUES. **Figure S6:** Schema of the algorithm behind HAPI.

**Supplementary Tables 1-6, SI_CCR5delta32_Tables.xlsx**

**Table S1:** CCR5delta32 proxy variants with an r² > 0.8, in the five EUR KGP3 populations. The table includes multiple sheets (**A-E**).

**Table S2:** Sample ID (1KGP3) for genomes identified with CCR5delta32, Haplotype A, B, and C, and their homologous recombinations. The table includes multiple sheets (**A-F**) and related to **Figure 1B** and **2A**.

**Table S3:** Inquiry from CCR5delta32 and Haplotype A’s tag SNPs in the GWAS catalog and Phenoscanner database. The table includes two sheets (**A-B**).

**Table S4A:** Detailed view of ancient genomes as classified by the HAPI model and the curated filters. The table includes multiple sheets (**A-C**).

**Table S5:** Summary of parameter estimates obtained using CLUES.

**Table S6:** Parameter estimates obtained using the Spatio-Temporal diffusion model.

## Materials and Methods

### Data

The modern dataset is constituted of whole-genome sequencing data of 2,535 individuals from 26 populations which were generated by the 1000 Genomes Project Phase 3 (1KGP3, http://www.1000genomes.org/), assigned to the following 5 super populations: African (AFR), admixed from the Americas (AMR), East Asian (EAS), South Asian (SAS), and European (EUR)^94^.

The ancient dataset comprises a total of 860 shotgun-sequenced genomes from various regions across Eurasia. The dataset includes genomes from the Stone Age^77^ (NEO samples ID, age: 11,000-3,000 BP), the Bronze Age^76^ (RISE samples ID age: 11,000-3,000 BP), and the Viking Age^75^ (VK samples ID, age: 2450 BP - CE 1600). The dataset for the Stone Age targets the Mesolithic and Neolithic Age and includes 317 genomes from archaeological sites across Europe. The sampling collection was particularly dense in northern Europe, involving 100 samples from Denmark (ENA Project ID: to be published soon). For the Bronze Age, 101 genomes were included from archaeological sites across Europe and Central Asia (ENA Project ID: PRJEB9021). Finally, the Viking Age dataset consisted of 442 genomes obtained from archaeological sites across Europe and Greenland, with a dense collection in Northern Europe (ENA Project ID: PRJEB37976).

### Identification of the CCR5delta32 deletion and the haplotypes

We used the LDLink 3.0 web tool, which includes the LDmatrix and LDpair modules^95^, to identify the CCR5delta32 proxy SNPs within the European (EUR) population of the 1KGP3 dataset (Table S1). These results were then explored in additional 1KGP3 populations. The haplotypes were called with samtools mpileup^96^ (**Table S2**), by using the region (chr3:46200000-46800000) from all available 1KGP3 whole-genome bam files. To determine the effect of the 86 tag SNPs belonging to Haplotype A, we employed Ensembl Variant Effect Predictor (VEP)^65^, while the GWAS catalog ^67^ and PhenoScanner V2 ^71,72^ were used to evaluate possible genotype-phenotype associations of tag SNPs (**Table S3**). All annotations refer to the human reference genome GRCh37 assembly.

### Development of the Haplotype-Aware Probabilistic model for Indels (HAPI) Simulations

We used gargammel (v. 1.1.2)^97^ to simulate a total of 144 ancient genomes at 8 different coverages (0.3X, 0.5X, 0.8X, 1X, 1.5X, 2X, 5X, 10X), using empirical read length distributions and post-mortem damage derived from 6 real ancient genomes (NEO78, NEO79, NEO752, VK287, VK543, VK526) from our dataset (**Figure S2A**). We simulated 48 genomes for each genotype (RR, RD, DD) using combinations of two versions of the GRCh37 human genome reference: a canonical, and one in which we manually added the CCR5delta32 deletion and the 86 variants from the haplotype (here referred to as “collapsed reference”) using the tool FastaAlternateReferenceMa er from GATK (v.4.1.8.1).^74^ The reads were simulated from HiSeq 2500 Illumina single-end runs with a length of 81 base pairs including adapters.

### Processing of simulated and ancient genomes

For the simulated genomes we used AdapterRemoval (v.2.1.3)^98^ with parameters “--mm 3 -- minlength 30 --min uality 2” to trim the reads from the simulated genomes at a length of at least 30 bp and to remove bases with quality 2 or less. We used bwa aln (v.7.16a)^99^ to map the adaptor-trimmed reads to both the canonical and the collapsed human reference genome (GRCh37) with seed disabled (parameter “-l 1024”) to allow for higher sensitivity in ancient DNA.^100^ We sorted the resulting alignments with samtools (v.1.9)^101^, removed duplicates with Picard MarkDuplicates (v1.128)^102^ and realigned the reads using GATK (v3.3.0)^74^ with Mills and 1000G gold-standard insertions and deletions. Finally, the alignment files were converted to cram with samtools view and indexed with samtools index. The ancient genomes were aligned to both the canonical and the collapsed human genome reference using the same pipeline as the simulated genomes except that the read groups were first merged to the library level, then duplicates were removed using Picard (v.1.128)^102^, and then the files were merged to sample level. Sample level bam files were subsequently realigned using GATK (v.3.3.0)^74^ and then converted and indexed to cram format. The workflows were implemented using Snakemake (v.5.12.0).^103^

### Processing of the 15 human genomes from the 1KGP3 dataset

We used 15 genomes from the 1KGP3 to benchmark the model, 5 for each deletion genotype: 5 RR (HG00179, HG00185, HG01500, HG01510, HG00159), 5 RD (HG00171, HG00267, HG01537, HG01605, HG00264), and 5 DD (HG00320, HG00323, HG01684, HG01762, HG00137). The genomes were aligned to both the canonical and the collapsed human genome reference with the same pipeline used for the alignment of the ancient genomes.

### Haplotype-Aware Probabilistic model for Indels (HAPI)

We developed a probabilistic model to combine the information from the 4 variants in the highest pairwise LD with the deletion (rs113341849, rs113010081, rs11574435, and rs79815064, r^2^ > 0.90, CEU) as a Prior, and the information from the reads mapping the CCR5delta32 deletion region as a Likelihood. The variants were called using samtools mpileup as implemented in pysam (v.0.16.0.1) in python. For a detailed overview of the algorithm see **Figure S6.** For each sample, we calculated the posterior probability for each deletion genotype (*RR*, *RD*, *DD*) as

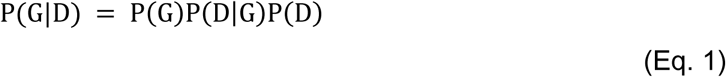

where *P*(*G*) is the prior probability of the given deletion genotype calculated using the information from the 4 variants (see eq. 2 below), *P*(*D*|*G*) is the likelihood of the deletion genotype based on the reads mapping to the CCR5delta32 deletion region (either canonical or collapsed reference) (see eq. 3 below) and *P*(*D*) is the marginal probability of the data. For the prior, we calculated the posterior probability of each deletion genotype using a simple bayesian genotyper based on the one developed by Mckenna et al, 2010^104^ as

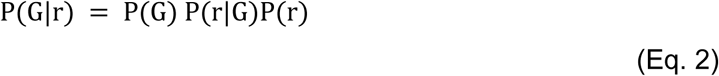

where *G* is the given SNP genotype (*ref*|*ref*, *ref*, *alt*, *or alt*|*alt*) and *r* is the data (the read base pileups mapping to each variant). We assume a uniform prior distribution for *P*(*G*), *P*(*r*) is the marginal probability of the data, and *p*(*r*|*G*) = *p*(*b*|*G*), where b represents each base covering the target locus. The probability of each base given the SNP genotype, considering only alleles from the reference and deletion genotype, is defined as

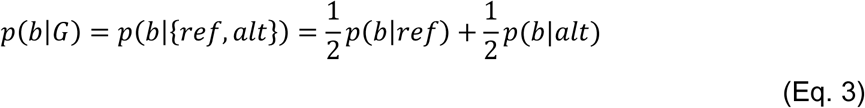

when the genotype G is decomposed into its two alleles. For simplicity, here we assumed that a sample having the genotype RR, RD, or DD also carries each of the four variants in the SNP genotype ref|ref, ref|alt, or alt|alt, respectively. The probability of observing a base given an allele is

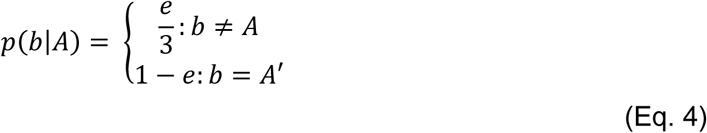

where *e* is the reversed *phred* scaled quality score at the base. At this point, each of the four variants has a posterior probability *P*(*G*|*r*) for each deletion genotype (RR, RD, DD). We scaled the posterior of each variant by the LD r^2^ value it has to the deletion in the CEU population. For each deletion genotype, we calculated the prior of eq. 1 as the joint probability (calculated with the specific multiplication rule, assuming each variant to be independent for simplicity) of the posteriors of the four variants, and we finally normalized them between 0 and 1 (subtracting by max and dividing by the sum). To calculate the Likelihood, we mapped the reads of each sample against two reference genomes: the canonical, and the collapsed one. The reads mapping to the canonical and collapsed references, together with their minimum overlapping lengths δ, were used to compute the Likelihood of each deletion genotype *RR*, *RD*, *DD* as follows. For each of the two references, and for each read, we calculated the probability of observing the read given the specific reference with

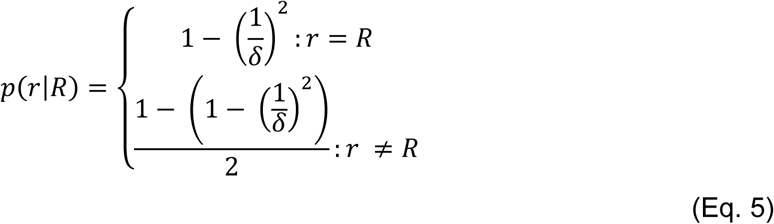

an adaptation from^104^, where *R* is the specific reference used for the mapping, i.e. canonical or collapsed. We then calculated the probability of observing the reads given the deletion genotype with

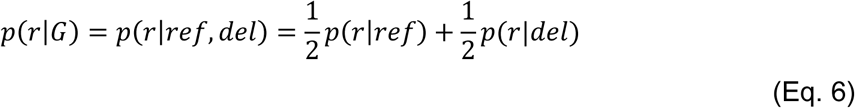

The genotype likelihood for each reference was then calculated with *p*(*D*|*G*) = *p*(*r*|*G*). The final genotype likelihood for each deletion genotype was computed as the joint probability of the likelihoods for the individual references (canonical and collapsed) with *p*(*D*|*G*) = *p*(*D*|*G*). Finally, for each sample, HAPI outputs three posterior probabilities for each deletion genotype RR, RD, DD, summing up to 1.

### Determining minimum overlapping length

We assigned a minimum overlapping length (δ) to each read mapping the deletion region, either on the canonical or collapsed reference. The δ represents the minimum number of nucleotides the reads overlap either the 5’ or the 3’ of the locus coordinates (See **Figure S2B** for a detailed clarification example). The CCR5delta32 has 4 equivalent representations, each with its own coordinates (https://varsome.com/variant/hg19/rs333?annotation-mode=germline). Thus, for each read mapping to the canonical reference, we calculated its minimum overlapping length δ by averaging across the δs calculated for each of the representations’ coordinates. A value of δ = 32 was assigned to the reads overlapping both the starting and ending coordinates of the canonical reference. For the collapsed reference, we calculated δ based on the coordinate 3:46414943 (GRCh37). For all the reads mapped, only those having a value of δ equal or greater than 6 were kept. The reads mapping to both deletion regions (from the canonical and collapsed references) were assigned to the reference to which they mapped with the lowest number of mismatches. This was done because, during the alignment of the simulated ancient DNA genomes, we observed that reads originating from the canonical deletion region mapped to the collapsed deletion region with a significantly higher number of mismatches compared to when they mapped to the canonical deletion region (and vice-versa) (signed test, p-value < 0.0001) (**Figure S3B).** Reads mapping to both references with the same number of mismatches were assigned to the reference to which they mapped with the highest δ. Reads mapped to both references with same number of mismatches and the same δ were discarded. The read mappings were analyzed using pysam (v. 0.16.0.1).

### Optimizing the model

During the developmental stage, we explored different approaches to optimize the model. To investigate how the minimum overlapping length of the reads across the deletion region influences the performance of HAPI, we ran the model using 10 different δ thresholds, from 1 to 10, on the simulated data. As expected, increasing the δ threshold resulted in an increase in the performance of the model from an MCC of 0.75 with δ=1 to a value of 0.873 with δ=10 (**Figure S3C**), but at the expense of having less reads satisfying the threshold and thus less genomes recovered (121 with δ=1 and 107 with δ=10) having at least one read mapping to the deletion region. We arbitrarily selected the δ threshold of 6 (corresponding to 6 nucleotides flanking each side of the breakpoint) because we found it to be a good compromise between performance and the number of genomes recovered (MCC = 0.81, genomes recovered 116). Additionally, we investigated rescaling the bam files to account for DNA damage and excluding reads without a perfect match in the alignment. Here, we found that rescaling did not have any significant effect on the performance of the model and that using only perfect match reads improved the performance of the model but at the expense of losing 22 genomes. These strategies were therefore not included in the final model.

### Applying the model to ancient genomes

To be analyzed by the model, a genome must have at least one read mapping to the CCR5delta32 deletion region with a minimum overlapping length δ of 6. The model was run on the simulated, ancient, and 1KGP3 genomes and we classified the genomes as being RR, RD, DD based on the highest posterior probability among the three, with a classification threshold of 0.5. To take into account the fact that the flanking regions of the deletion include repeated nucleotides, and that two of the 4 variants used for calculating the haplotype-informed prior look like ancient DNA damage, we applied two manual curation steps. In the first one “permissive filter” we manually re-classified some genomes if they had only 1 SNP called, and the same SNP looked like aDNA damage (G to A, and C to T). In the second one “strict filter”, we re-classified the genomes which we think have Haplotype B instead of the deletion, because of no reads covering the deletion but only the reference.

### Benchmarking using Haplotype Caller

The 15 genomes selected from the 1KGP3 population were processed using HaplotypeCaller from GATK (v. 4.1.9.0)^74^ to produce vcf files with SNP and indels calls using the following options: --intervals 3:46277577-46457412 --interval-padding 100 --stand-call-conf 30.0 -ERC BP_RESOLUTION. The vcf files were left aligned and normalized using bcftools norm (v. 1.10.2^105^ and then processed in R (v.4.0.3)^106^.

### Local ancestry of individuals harboring CCR5delta32 deletion in the 1KGP3

We used HaploNet^73^ on the full 1KGP3 dataset to generate haplotype cluster likelihoods in windows along the genome with default parameters of “haplonet train” besides “--x_dim 512”, such that the genomic windows had a fixed size of 512 SNPs. e used the haplotype cluster likelihoods to estimate ancestry proportions with an assumption of K=5 ancestral populations, representing the 5 super populations of 1KGP3, using the “haplonet admix” command. The haplotype cluster likelihoods and ancestry proportions were then finally used to infer local ancestry for all genomic windows in the individuals with the CCR5delta32 locus using the “haplonet fatash” command.

### CLUES analysis

To reconstruct the allele frequency trajectory of the CCR5delta32 deletion, we used a modified version of the software CLUES, adapted for time-series data^80,81^. We converted the output of HAPI into hard called genotypes, using the outputs from the permissive and strict filters. We then conditioned the inference of the trajectories on a present-day frequency of 0.1237 and an estimate of the effective population size history, inferred from genomes in the Finnish (FIN), British (GBR), and Tuscan (TSI) populations from the 1KGP3^94^, using the software Relate^107^. The code to reproduce these analyses is available in the Github repository https://github.com/ekirving/ccr5_paper.

### Estimating the age of CCR5delta32

To infer an estimate for the age of CCR5delta32, we extracted the time series of posterior probability densities from all the CLUES models. As CLUES does not have an explicit mutational model, we approximated the temporal origin of the CCR5delta32 mutation by finding the most recent time-point in which the majority of the posterior density was assigned to the two lowest frequency bins – i.e., the time point at which the model estimates that there is a greater than 50% probability that the allele is at the lower limit of possible frequency values. For each genotype call set, we averaged the approximated allele ages inferred from CLUES in the models with and without conditioning on the present-day frequency from 1KGP3, and used the resulting average as an input parameter for the spatiotemporal model.

### Method for modeling the spatiotemporal diffusion of the deletion allele

To model the diffusion of the CCR5delta32 allele across space and time, we use a method described in Muktupavela et al. 2021^82^ and available from: https://github.com/RasaMukti/stepadna. We adapted the method so that the input genotype calls for each individual corresponded to the genotype with the highest posterior probability obtained from HAPI. To do this, we modified the equation (5) from^82^:

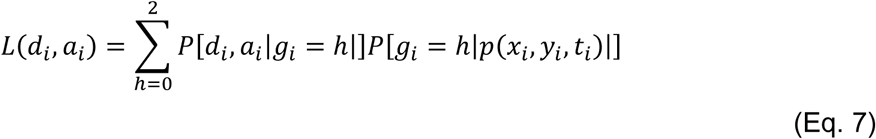

Here *L* is the likelihood of the observed data for individual *i*, *a*_*i*_ and *d*_*i*_ represent the number of reads carrying ancestral or derived alleles, respectively, *g*_*i*_ ∈ {0,1,2} is the genotype of the individual at the particular locus, (*x*_*i*_, *y*_*i*_) represent the coordinates of the sampling location for that individual and *t*_*i*_ is the estimated sample age. *P*[*d*_*i*_, *a*_*i*_|*g*_*i*_ = ℎ|] is the likelihood for genotype *g*_*i*_ and *P*[*g*_*i*_ = ℎ|*p*(*x*_*i*_, *y*_*i*_, *t*_*i*_)|] corresponds to binomial distribution, where *p*(*x*_*i*_, *y*_*i*_, *t*_*i*_) is the solution to a reaction-diffusion partial differential equation and it represents the allele frequency distribution across a two-dimensional (*x*, *y*) landscape at a time point *t* :

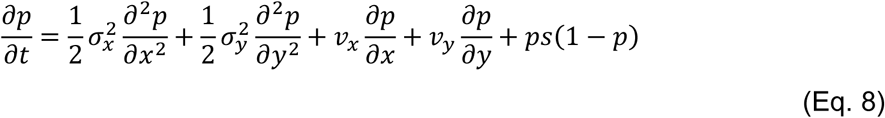

where *σ*_*x*_, *σ*_*y*_ are the longitudinal and latitudinal diffusion coefficients, respectively, *v*_*x*_ and *v*_*y*_ represent the longitudinal and latitudinal advection coefficients, respectively, and *s* is the selection coefficient. We modified the equation so that the likelihood of the genotype *g*_*i*_ is equal to 1 if the genotype corresponds to the genotype with the highest posterior probability and 0 otherwise.

We applied the method to the different deletion call datasets, combining them with the present-day geographically-spread deletion calls compiled in Novembre et al. 2005^12^ (**Figure S4**). We removed genomes that were outside of the geographic area bounded latitudinally by 30°N and 75°N and longitudinally by 10°W and 80°E.

Maximum likelihood optimization was carried out by initializing 50 points in the multi-parameter space and using a first round of simulated annealing^108^ followed by a run of the L-BFGS-B algorithm^109^ to refine the optimization.

### Code availability

The simulated sequence data mapped to the GRCh37 and to the Collapsed reference, restricted to the CCR5Delta32 region, are available at https://doi.org/10.17894/ucph.a31d9052-546d-4f8f-8e16-e5bd896df67b together with the results of running HAPI on them. The HAPI model is available as a pip package at https://pypi.org/project/hapi-pyth/ and instructions on how to install and run it are available at https://github.com/RasmussenLab/HAPI. The code for the CLUES analysis is available at https://github.com/ekirving/ccr5_paper. The code for reproducing the spatiotemporal diffusion analysis can be found at https://github.com/RasaMukti/ccr5delta32_analysis. Any additional information required to reanalyze the data reported in this paper is available from the lead contact upon request.

