## Supplementary Figures 1-6 for "Tracing the evolutionary path of the CCR5delta32 deletion via ancient and modern genomes"

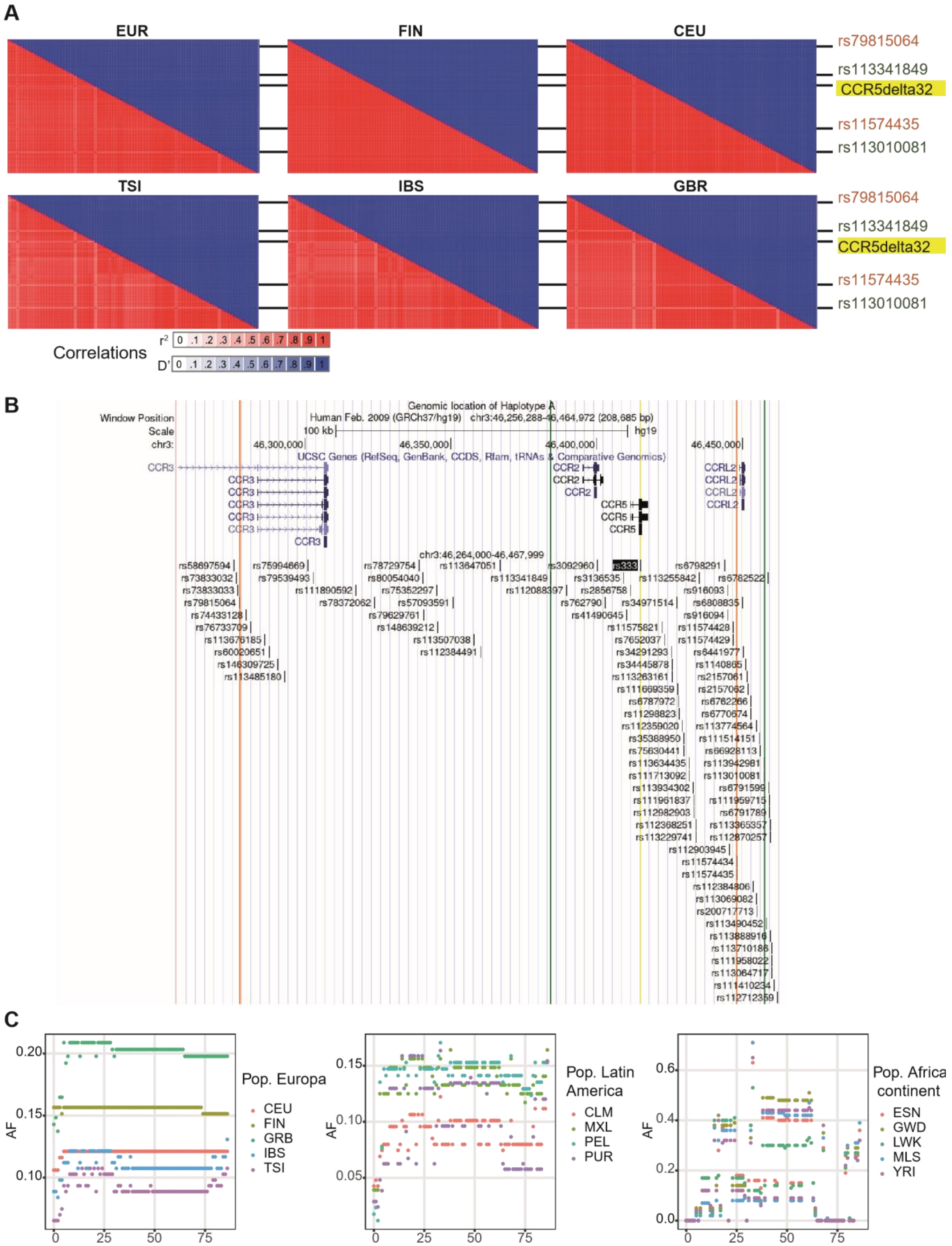

**Figure S1. Details information of Haplotype A: LD statistics, genomic location and the AF of the proxy variants.**

**A)** Heatmap matrix of pairwise LD statistics from Haplotype A in the five EUR populations: EUR (FIN, CEU, TSI, IBS, and GBR) followed by each EUR population separately: $r^2$ values are in shades of red, while D' values are in shades of blue, wherewith darker colors indicate a higher degree of LD. The strong LD pattern from Haplotype A is observed in the FIN and CEU populations, whereas the pattern becomes weaker in southern and western Europe. The weaker LD patterns are caused by the homologous recombinations of Haplotype A and the more frequent presence of Haplotype B and C and their homologous recombinations. **B)** The UCSC genome browser (https://genome.ucsc.edu) displays the location of Haplotype A on 3p21.31. The CCR5delta32 allele (rs333) is highlighted in yellow, and the two SNPs with $r^2 = 1$ are marked in green, while the two SNPs with $r^2 = 0.903$ are marked in orange. The haplotypes span ≈ 0.19 Mb and encompass *CCR3*, *CCR2*, *CCR5*, and *CCRL2*. Detailed information on the tag variants is included in Table S1. **C)** AF of CCR5delta32 and the 86 tag variants, in different countries and continents. The x-axis corresponds to Haplotype A, position 0 = CCR5delta32, and the 86 tag variants are ranked according to their r2 values. The y-axis shows the AF subtracted from the 1KGP3. Populations from Europe and Latin America all have individuals carrying the CCR5delta32 allele/Haplotype A, B, and C, whereas none of the individuals from the African continent carried any of the three haplotypes. However, precursor variants for Haplotype C exist, where 38 of the variants have a higher AF in the African population compared to the European population.

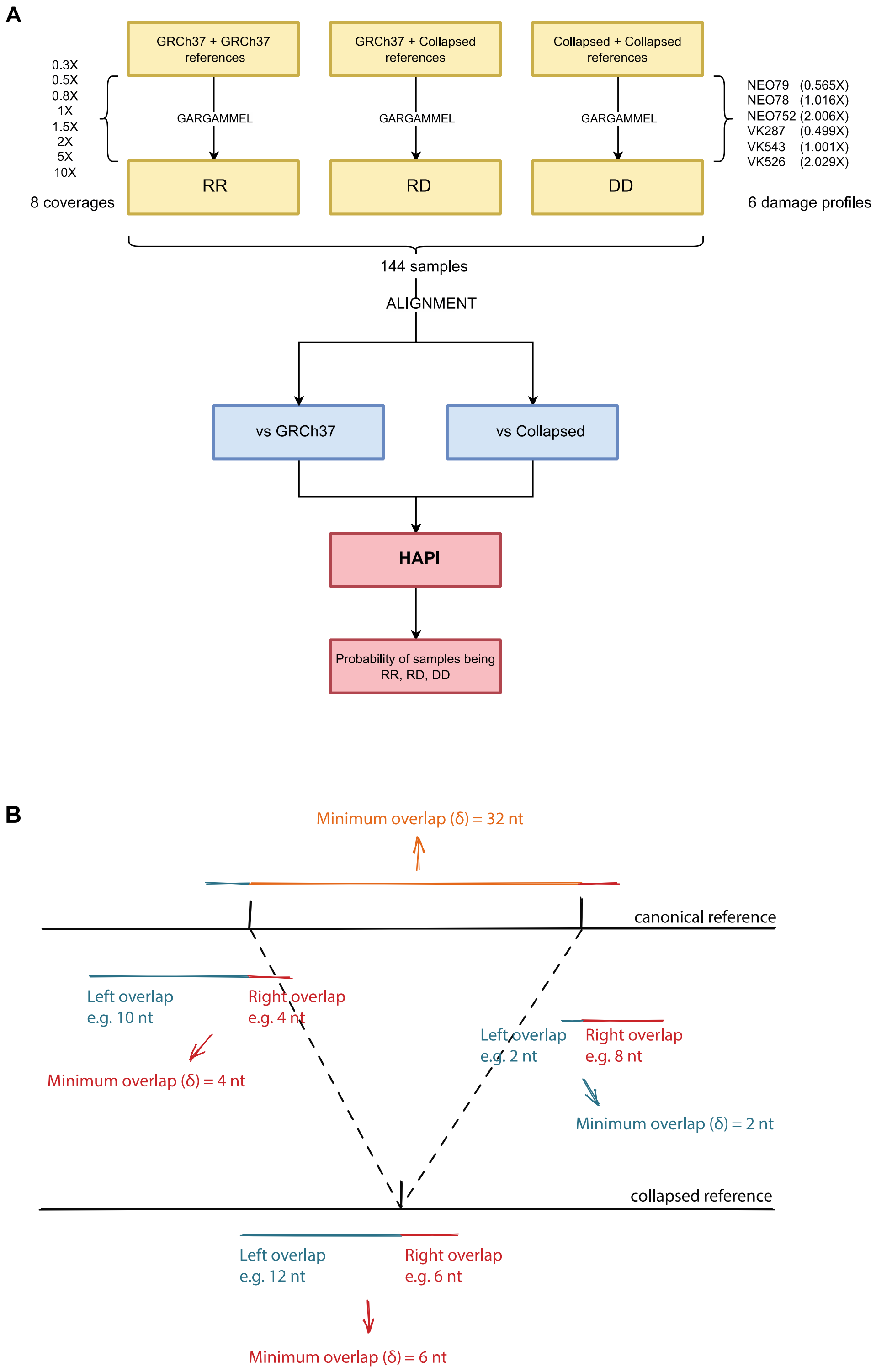


**Figure S2. Workflow of the data analysis on the simulated ancient samples and details of the overlapping lengths.**

**A)** The software Gargammel was used to simulate a total of 144 ancient samples using damage profiles derived from 6 real ancient samples (right), at 8 different coverages (left). After the simulation, the reads were aligned to the canonical GRCh37 and to the Collapsed reference using bwa. Finally, HAPI was used to calculate the probability of each sample having each of the three deletion genotypes (RR, RD, DD). **B)** Schema of the reads mapping to the two references. Each read mapping to the canonical and the collapsed references is assigned a minimum overlapping length δ, which represent the minimum number of nucleotides with which it overlaps the deletion coordinates. In order for a genome to be analyzed by HAPI, it needs to have at least one read mapping either the canonical or collapsed reference, with a minimum overlapping length of 6.

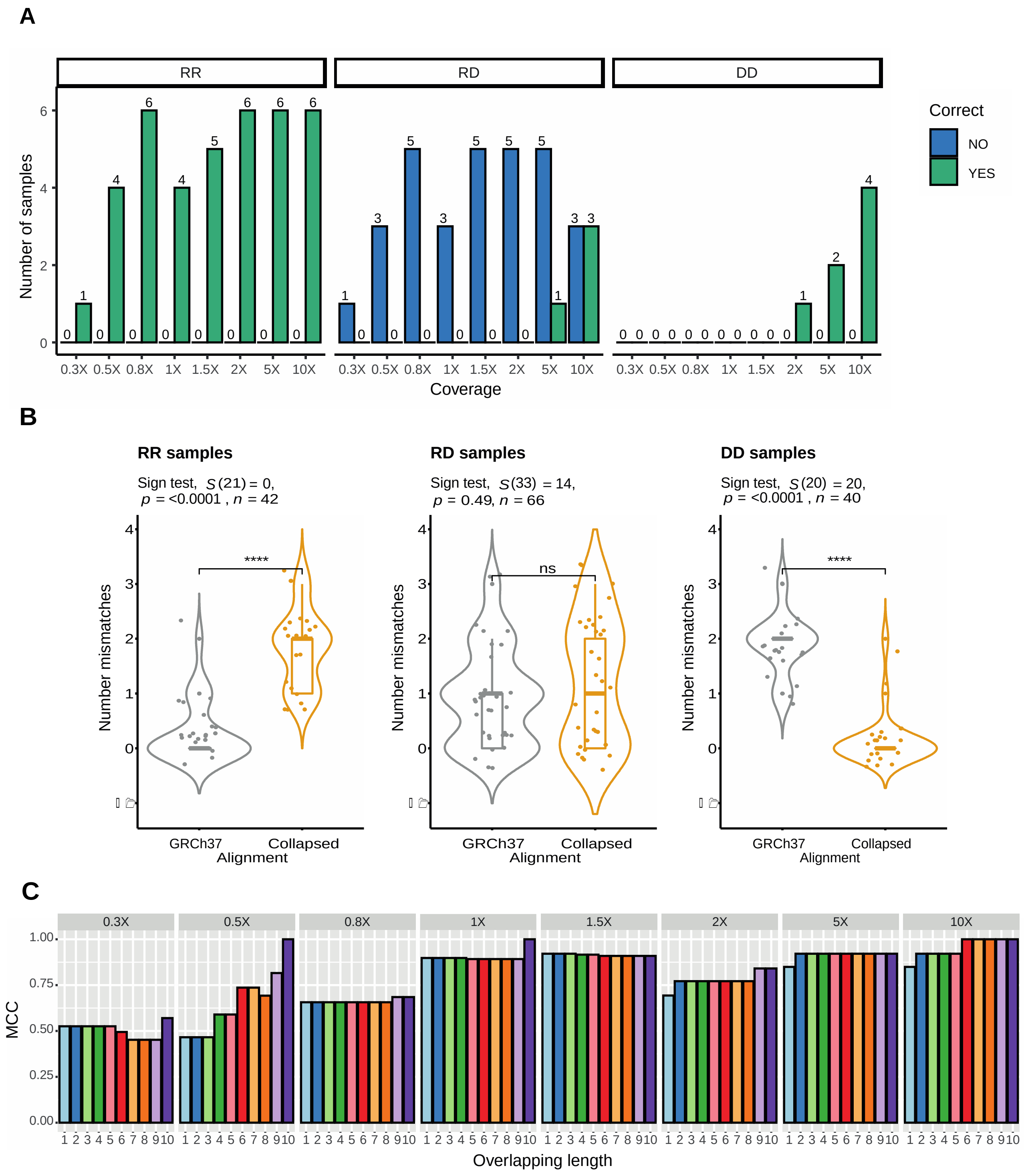


**Figure S3: Assessing GATK HaplotypeCaller, Mismatch Rates, and HAPI performance at different overlapping lengths**

**A)** Number of simulated samples classified by GATK HaplotypeCaller. Bars represent the number of ancient simulated samples correctly (green) or incorrectly (blue) classified by GATK

HaplotypeCaller, stratified by coverage (from 0.3 to 10X) and by deletion genotype (RR, RD, DD). A considerable number of samples were not classified by GATK HaplotypeCaller as it failed to detect any reads, thus their columns are 0. **B)** Results of the sign test for the number of mismatches of reads originating from simulated individuals carrying the RR, RD, and DD Deletion Genotypes when aligned to the canonical reference (GRCh37) or the collapsed reference (Collapsed). We can see that reads originating from individuals with DD genotype, and thus having the deletion, mapped to the reference genome with a higher number of mismatches compared to the collapsed reference (plot on the right). The opposite effect was seen in the plot on the left, while no significant difference was shown for reads originating from simulated individuals with RD genotype. **C)** MCCs of HAPI at different values for the parameter “overlapping length”. The performance of HAPI in MCC is shown at different values of the minimum overlapping length for the reads mapping to the deletion region. The MCC increases with higher overlapping lengths values, but at the expense of having less reads satisfying the requirements and thus less samples analyzed. A minimum overlapping length value of 6 was chosen (see Methods).

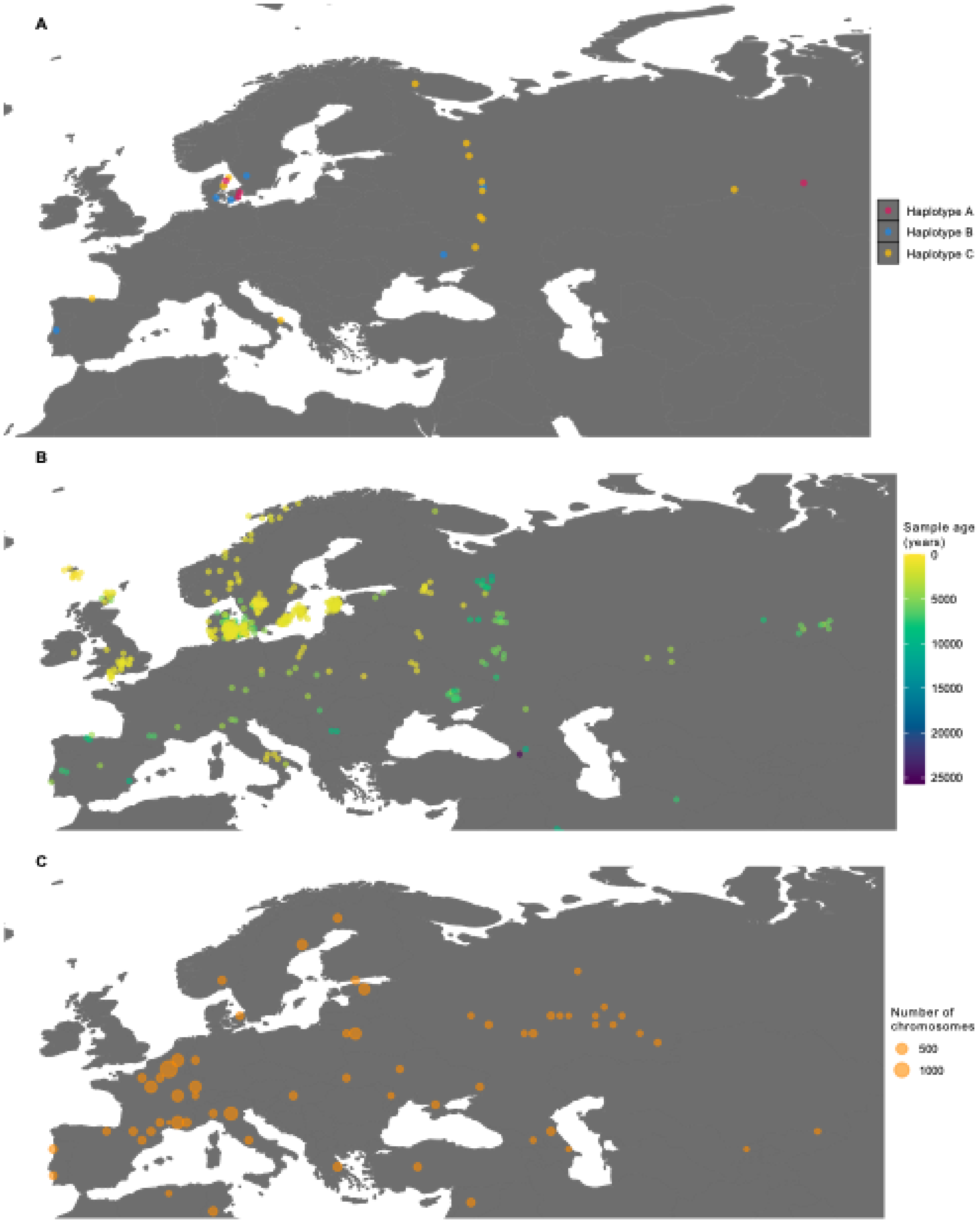


**Figure S4: Ancient and present sample distribution**

**A)** Neolithic samples carrying either haplotype A after applying the strict filter, or traces of either haplotype B or haplotype C. **B)** The ancient DNA sampling locations of samples used in the spatiotemporal analysis [1–3]. **C)** Present-day samples used in the spatiotemporal analysis. The data is compiled in Novembre et al. 2005[4].

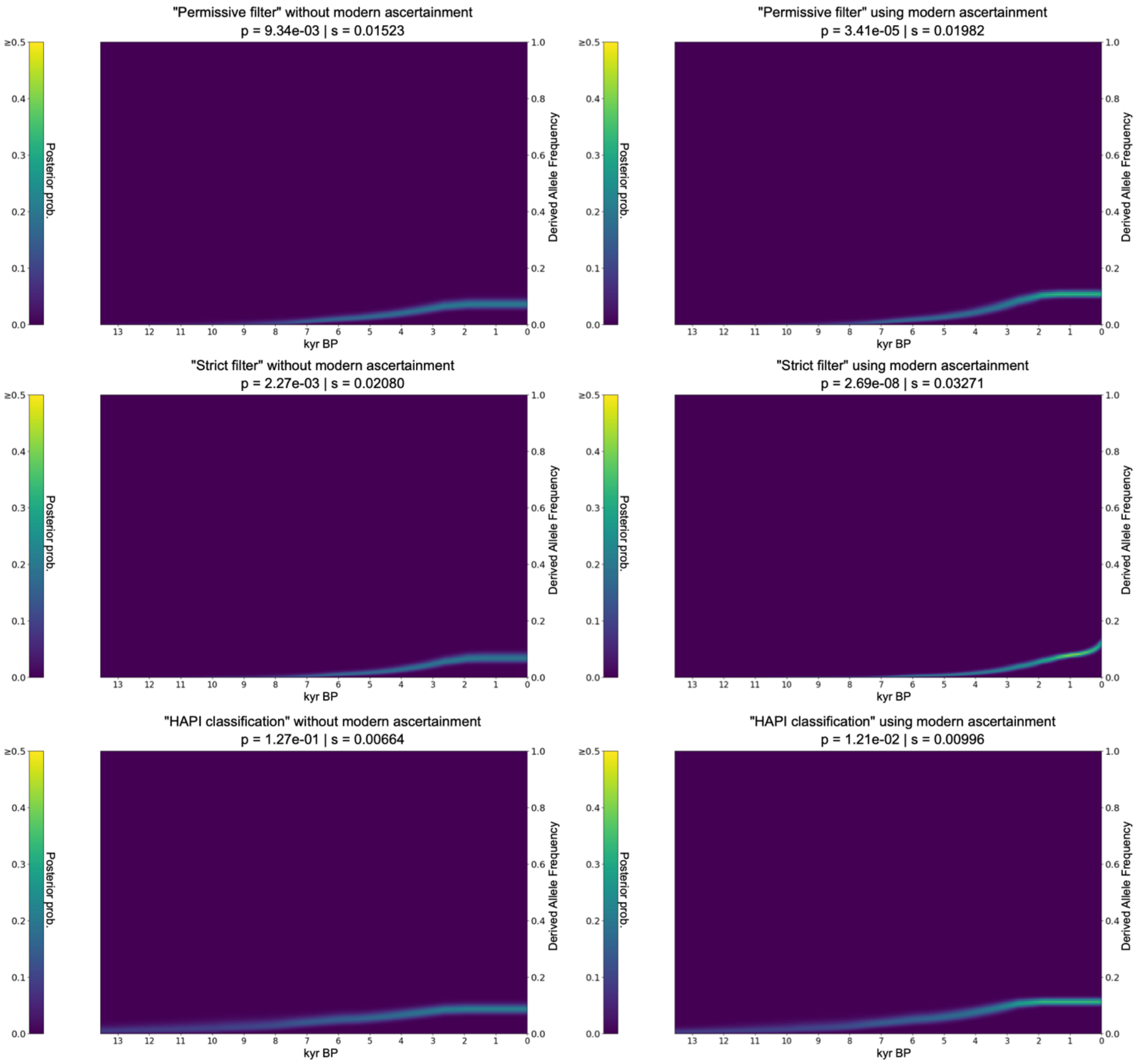


### Figure S5: Allele frequency trajectory inferred by CLUES

Allele frequency trajectory inferred by CLUES. Upper row shows results using permissive filter genotype call set, middle – using strict filter and bottom row shows results using HAPI classifications. Left column shows results using ancient data only and the right column corresponds to results when ancient data is combined with modern ascertainment from 1KGP3. In each figure the line represents posterior probability density. The p-value indicates evidence for rejecting a neutral model and we also provide the most likely selection coefficient inferred by CLUES.

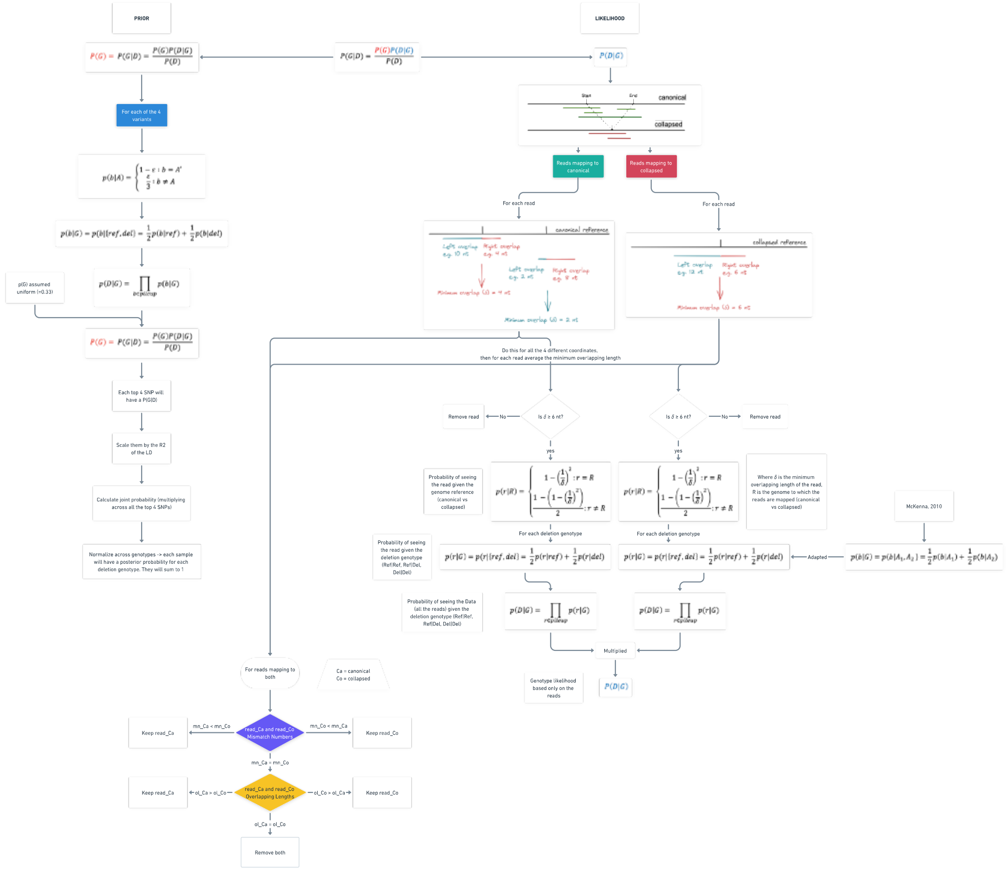




### Figure S6: Schema of the algorithm behind HAPI

The equation in the top middle represents P(G|D), i.e. the posterior probability for a sample having the deletion genotype RR, RD, or DD. The steps used to calculate the Prior are outlined on the left. The algorithm which calculates the Likelihood is illustrated on the right, and it takes into account how many reads map to either the canonical or the collapsed reference, as well as the overlapping lengths and the number of mismatches with which they align. More details are provided in the Methods.
